# Noninvasive molecular subtyping of pediatric low-grade glioma with self-supervised transfer learning

**DOI:** 10.1101/2023.08.04.23293673

**Authors:** Divyanshu Tak, Zezhong Ye, Anna Zapaishchykova, Yining Zha, Aidan Boyd, Sridhar Vajapeyam, Rishi Chopra, Hasaan Hayat, Sanjay Prabhu, Kevin X. Liu, Hesham Elhalawani, Ali Nabavizadeh, Ariana Familiar, Adam Resnick, Sabine Mueller, Hugo J.W.L. Aerts, Pratiti Bandopadhayay, Keith Ligon, Daphne Haas-Kogan, Tina Poussaint, Benjamin H. Kann

## Abstract

**Key Results:**

- An innovative training approach combining self-supervision and transfer learning (“TransferX”) is developed to boost model performance in a low data setting;
- TransferX enables the development of a scan-to-prediction pipeline for pediatric LGG mutational status (BRAF V600E, fusion, or wildtype) with ≥75% accuracy on internal and external validation;
- An evaluation metric, “COMDist”, is introduced to increase interpretability and quantify the accuracy of the model’s attention around the tumor.

**Purpose:** To develop and externally validate a scan-to-prediction deep-learning pipeline for noninvasive, MRI-based BRAF mutational status classification for pLGG.

**Materials and Methods:** We conducted a retrospective study of two pLGG datasets with linked genomic and diagnostic T2-weighted MRI of patients: BCH (development dataset, n=214 [60 (28%) BRAF fusion, 50 (23%) BRAF V600E, 104 (49%) wild-type), and Child Brain Tumor Network (CBTN) (external validation, n=112 [60 (53%) BRAF-Fusion, 17 (15%) BRAF-V600E, 35 (32%) wild-type]). We developed a deep learning pipeline to classify BRAF mutational status (V600E vs. fusion vs. wild-type) via a two-stage process: 1) 3D tumor segmentation and extraction of axial tumor images, and 2) slice-wise, deep learning-based classification of mutational status. We investigated knowledge-transfer and self-supervised approaches to prevent model overfitting with a primary endpoint of the area under the receiver operating characteristic curve (AUC). To enhance model interpretability, we developed a novel metric, COMDist, that quantifies the accuracy of model attention around the tumor.

**Results:** A combination of transfer learning from a pretrained medical imaging-specific network and self-supervised label cross-training (TransferX) coupled with consensus logic yielded the highest macro-average AUC (0.82 [95% CI: 0.70-0.90]) and accuracy (77%) on internal validation, with an AUC improvement of +17.7% and a COMDist improvement of +6.4% versus training from scratch. On external validation, the TransferX model yielded AUC (0.73 [95% CI 0.68-0.88]) and accuracy (75%).

**Conclusion:** Transfer learning and self-supervised cross-training improved classification performance and generalizability for noninvasive pLGG mutational status prediction in a limited data scenario.

## INTRODUCTION

Pediatric low-grade gliomas (pLGG) are the most common pediatric brain tumors, comprising up to 40% of tumors in this population (1). These tumors exhibit diverse clinical outcomes and molecular characteristics, often driven by an activating BRAF mutation, either the BRAF-V600E point mutation or fusion events. Molecular classification and segregation of wildtype tumors from BRAF subtypes is vital for accurate treatment selection and risk stratification in pLGG, particularly given the emergence of novel BRAF-directed therapies (2). The presence of the BRAF-V600E mutation, found in 15-20% of cases, was historically associated with poor survival, particularly when combined with CDKN2A deletion (3), though with targeted BRAF pathway-directed therapies this may be changing. BRAF-V600E mutated pLGG also exhibit an increased risk of malignant transformation while patients with BRAF fusion and neurofibromatosis type 1 have a favorable outcome (4). Accurate distinction between BRAF-V600E, BRAF-fusion, and wildtype tumors, plays a crucial role in determining prognosis and optimal treatment strategy.

Surgical resection for pLGG allows for assessment of mutational status. However, in over one-third of cases, resection, or even biopsy, may not be feasible nor recommended (5). In these situations, children may require alternative therapies to control a symptomatic tumor or undergo periodic MRI surveillance. Therefore, non-invasive imaging-based tumor molecular subtyping, if accurate and reliable, could enable proper selection of patients for BRAF-targeted therapies and clinical trials. In recent years, deep learning (DL) has emerged as the forefront technology for analyzing medical images (6,7), and has demonstrated numerous successful applications, encompassing tumor segmentation (8–10), outcome prediction (11,12), tumor and molecular classification (13,14). However, DL performance degrades dramatically in limited data scenarios, due to instability, overfitting, and shortcut learning (15), and a key barrier to applying DL to pLGG imaging, is the lack of training data available for these rare tumor cases. For these reasons, there has been limited success in using DL for pLGG mutational classification. Another barrier to clinical usability is that most algorithms require manual tumor segmentation as input, which is resource-intensive and requires specialized expertise. Few studies have been published investigating pLGG BRAF mutation classification using deep learning (16) and a combination of deep-learning and radiomics (17) but all of them present a single institution study and lack external validation.

Here, we address these gaps by developing and externally validating the first imaging based automated, scan-to-prediction DL pipeline capable of non-invasive BRAF mutational status prediction for pLGG. The pipeline comprises built-in pLGG segmentation, BRAF mutation classifiers, and a consensus decision block to predict BRAF mutation status. We leverage the pLGG dataset as our developmental dataset and a novel combination of in-domain transfer learning and self-supervision approach, called “TransferX” to maximize performance and generalizability in a limited data scenario. Additionally, to improve interpretability of our pipeline, we introduce a way to quantify the model attention via spatial maps, called Center of Mass Distance (COMDist) analysis. COMDist estimates the distance between the center of mass of the GradCAM heatmap and the tumor’s center of mass. Together, these methods enable practical, accurate, noninvasive mutational classification for pLGG.

## METHODS

### Study Design and Datasets

This study was conducted in accordance with the Declaration of Helsinki guidelines and following the approval of local Review Board (IRB). Waiver of consent was obtained from IRB prior to research initiation due to public datasets or retrospective study. This study involved two patient datasets: a developmental dataset from one high-volume academic institution (BCH; n=214), for training, internal validation, and hypothesis testing. This dataset included all children aged 1 – 25 with a tissue-confirmed diagnosis of WHO grade I-II glioma with BRAF mutational status information and available pretreatment T2-weighted (T2W) brain MRI seen at the institution from 1994 to 2022. A second data from the Children’s Brain Tumor Network (CBTN; n=112) was used for external validation. This dataset included all patients from the publicly available CBTN pLGG cohort who had T2W brain MRI and confirmed WHO grade I-II glioma tissue diagnosis and mutational status as above. BRAF status was determined by OncoPanel, which performs targeted exome-sequencing of 227 to 477 cancer-causing genes. BRAF mutational status may also have been captured by genomic sequencing via in-house PCR on tissue specimens. In cases where neither could not be performed, immunohistochemistry (IHC) was used to determine V600E status. BRAF-fusion status was determined by a gene fusion sequencing panel. DNA copy-number profiling via whole-genome microarray analysis was also performed in some cases. We report our results in accordance with the Checklist for Artificial Intelligence in Medical Imaging (CLAIM) guidelines (18). A portion of patients from the CBTN dataset (n=140) and an additional subset from the BCH dataset (n=100) had been utilized in two previous studies (10,19). It’s worth highlighting that these prior investigations were centered around tumor segmentation, whereas the present study was primarily dedicated to identifying BRAF mutational subtypes.

### Deep Learning Pipeline

The proposed pipeline for mutation class prediction operates in two stages (Fig. 1A). The initial stage involves T2W-MRI preprocessing (Supplemental Methods: SM 3, SM 2) and input to a nnUNet-based 3D tumor auto-segmentation model previously developed, externally validated, and clinically benchmarked by our group (pipeline available at: https://github.com/AIM-KannLab/pLGG_Segmentation) (10). This first stage outputs a preprocessed, skull-stripped image along with a corresponding segmentation tumor mask (Fig. 1B) (Supplemental Methods: SM 4).

**Figure 1.**
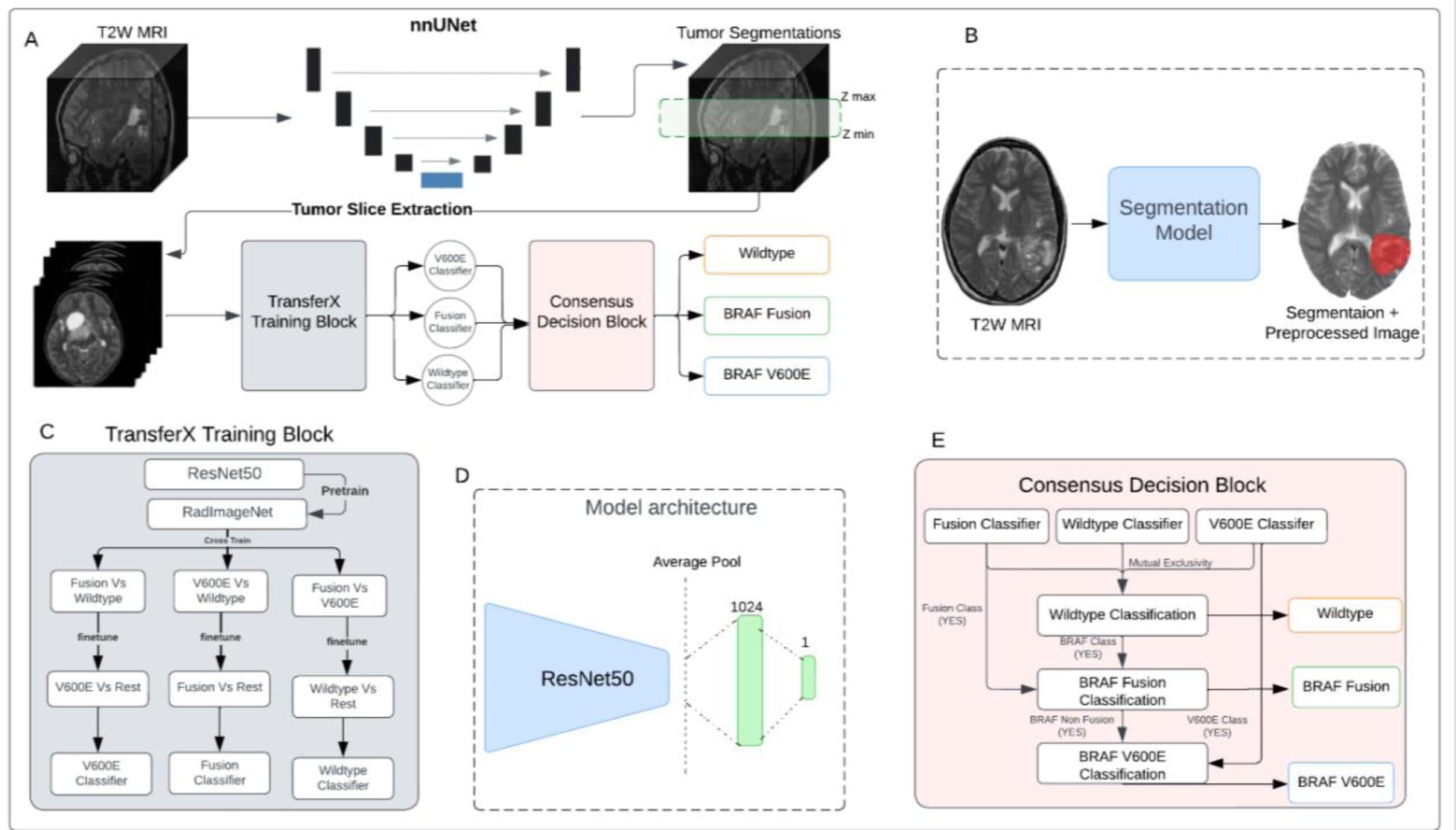
(A) Schematic of the scan-to-prediction pipeline for molecular subtype classification. The pipeline inputs the raw T2W MRI scan and outputs the mutation class prediction. (B) Input and output depiction of the segmentation model from stage 1 of the pipeline. The segmentation block also involves registration and preprocessing of the input scan. The output consists of the preprocessed input MRI scan along with the co-registered segmentation mask. (C) Flow diagram of the TransferX training block and approach. The TransferX algorithm is employed to train three individual subtype classifiers (BRAF-V600E, BRAF-Fusion, Wild-type). (D) The model architecture of individual binary molecular subtype classifier. (E) Schematic of consensus decision block. The block inputs the classification outputs and corresponding scores from the three individual subtype classifiers and fits them into a consensus logic, and outputs the final predictions. The mutational class predictions are output sequentially where the input is first checked for wild-type or non-BRAF class first. If the input doesn’t belong to wildtype or non-BRAF class, then the logic progresses to check the BRAF mutation class with BRAF-Fusion checked first then followed by BRAF-V600E. T2W: T2-weighted.

**Figure 2.**
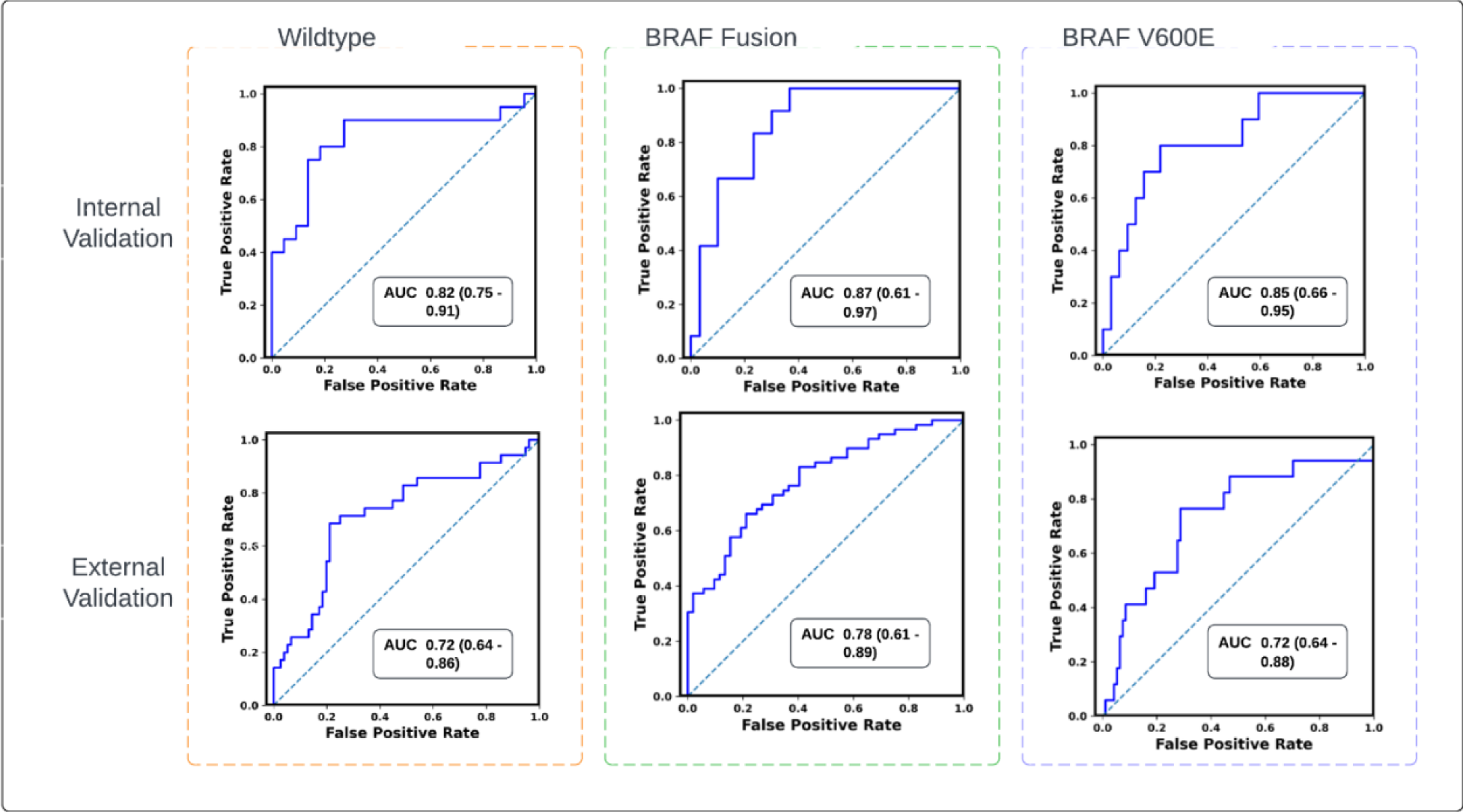
Receiver operating characteristics (ROC) curves of the scan-to-prediction pipeline’s predictions for all the three molecular subtype classes for internal validation (n=59) and external validation (n=112). The models, trained with TransferX, form the individual subtype classifiers. The outputs of the subtype classifiers are pooled using consensus logic, to result the pipeline predictions for each mutation class.

The second stage of the pipeline encompasses three binary subtype classifiers (BRAF-Fusion vs. rest; BRAF-V600E vs. rest; Wild-type vs. rest), each specifically trained to identify one of the following classes: BRAF-V600E, BRAF-Fusion, and Wild-type (Supplemental Methods: SM 5-6). For each subtype classifier a ResNet-50 model (20) was chosen as the fundamental encoder for extracting feature embeddings from 2D images, given its high performance on medical imaging classification problems (21,22) and the availability of pretrained network weights (23). The fully connected layers succeeding the average pooling layer of the ResNet-50 were replaced by a layer of 1024 neurons, and a final layer of single neurons for binary classification (Fig. 1D, Supplemental Methods: SM 6). Following binary classification from each binary subtype classifier, a consensus decision block collates the predictions from the classifiers, yielding the overall mutational status (Supplemental Methods: SM 7) (Fig. 1E). The final output of the consensus decision block and the pipeline consequently is a classification decision and its corresponding probability.

Three different strategies were investigated for training individual binary classifiers. The initial approach, training from scratch, involved initializing the binary classifier model with random weights. For the second approach, called RadImageNet Finetune, the classifier model was initiated with pretrained weights from the RadImageNet (23) for the ResNet-50 model. This prior initialization was intended to yield superior feature embeddings compared to random weight initialization and training from scratch or out-of-domain transfer learning (24).

### TransferX

The third approach, called TransferX, starts with pretrained weights from RadImageNet, but then adds two sequential stages of finetuning on separate, but related, classification tasks which act as pretext tasks for self-supervision, followed by a final finetuning on the target class (Fig. 1C). As an illustrative example, the training of a BRAF-fusion classifier began with initialization via pretrained RadImageNet weights and sequential finetuning for BRAF-V600E prediction, followed by Wild-type prediction, and finally finetuning for BRAF-Fusion prediction. We hypothesized that combining transfer learning and self-supervised cross-training would enable the model to learn stronger, more generalizable features for mutational status prediction by exposure to different, though similar, classification problems. The Models were trained to minimize loss at the axial slice-level on the development dataset and internally tested on an internal validation set (25% of data randomly selected; Supplemental Methods: SM 4, Fig. S3) and tested on the external validation dataset.

### Performance Evaluation and Statistical Analysis

Since each of the MRI scan of each patient was factored into multiple tumor slice images to generate aggregated patient-level prediction, the output probability scores of the individual 2D axial images were averaged to calculate the patient level probability score. The patient-level classification was then done by applying a threshold on the patient level probability score [Eq 1].

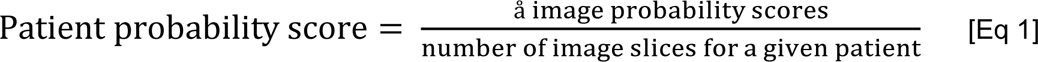

The primary performance endpoint was the area under the curve (AUC) of receiver operating characteristic (ROC) at the patient-level. We calculated composite AUC and accuracy based on a weighted average of the output of the three mutational subtype classifiers. The three DL approaches were initially evaluated on the internal test set, and the highest performing model was locked for external validation. Secondary endpoints included sensitivity and specificity, precision, and accuracy, and were calculated using the model output, thresholded to optimize the Youden Index (25) on the internal test set. Post-hoc calibration was applied on the internal validation set and model calibration was assessed graphically pre- and post-calibration (Supplemental Methods: SM 8; Fig. S7). We compared AUC’s for different models and calculated 95% Confidence Intervals (CIs) using the DeLong method (26). The standard error of the AUC was calculated considering the numbers of positive and negative cases in the sample, and the derived variance of AUC. A two-sided *p*-value of <0.05 was considered statistically significant. Statistical metrics and curves were calculated using Scikit-learn packages (27) in Python v3.8.

### Center of Mass Distance Analysis (COMDist) to evaluate model attention

To enable the use of Gradient-weighted Class Activation Maps (GradCAM) (28) as a quantitative performance evaluation tool, we developed “Center of Mass Distance” (COMDist), a quantifiable metric for comparing GradCAM images across different methodologies (Fig. 4C). COMDist calculates and averages the distance (in mm) between the tumor’s center of mass (from the segmentation mask) and the center of mass of the GradCAM heatmap over the entire dataset, with smaller values indicating that the model is more accurately focusing on the tumor region (Fig. 4B).

## RESULTS

### Patient Characteristics

The total pLGG patient cohort consisted of 326 pLGG patients from two cohorts, with 214 patients in the development set from BCH cohort and 112 patients in the external test set from CBTN (Table 1). Median age was 6 (range: 1-21) in the CBTN cohort and 5 (range: 1-20) in the BCH cohort. All patients had pathologically or clinically diagnosed grade I/II low-grade glioma, with a mixture of histologic subtypes and intracranial locations. The developmental dataset contained 50 (23%), 60 (28%), and 104 (49%) patients with BRAF-V600E, BRAF-Fusion, and Wild-type, respectively, and the external validation dataset contained 17 (15%), 60 (53%), and 35 (32%) patients with BRAF-V600E, BRAF-Fusion, Wild-type, respectively (Table 1). Age and sex were not associated with BRAF mutational status (Table S4, Fig. S5). Categorical variables of tumor locations were one hot encoded, and a logistic regression model was trained for each molecular subtype with an accuracy of 59%, 52%, 63% for BRAF V600E, BRAF Fusion, and Wild-type respectively, proving that tumor location can not be employe as the only variable to perform molecular subtype classification.

**Table 1.** Patient cohort characteristics.

|  | Development<br>(BCH, n = 214) | External Validation<br>(CBTN, n = 112) | p-values |
| --- | --- | --- | --- |
| **Age (years)** |  |  | 0.19* |
| median (range) | 5 (1 – 20) | 6 (1 - 21) |  |
| **Sex n (%)** |  |  | 0.82<sup>+</sup> |
| Female | 95 (44.4%) | 51 (45.5%) |  |
| Male | 113 (52.8%) | 55 (49.1%) |  |
| Unknown | 6 (2.8%) | 4 (3.6%) |  |
| **Race/Ethnicity n (%)** |  |  | 1.076e-06<sup>+</sup> |
| Non-Hispanic Caucasian/white | 145 (67.8%) | 71 (64.5%) |  |
| African American/Black | 6 (2.8%) | 14 (12.7%) |  |
| Hispanic/Latinx | 3 (1.4%) | 10 (9.1%) |  |
| Asian American/Asian | 9 (4.2%) | 3 (2.7%) |  |
| American Indian/Alaska Native | 0 | 1 (0.9%) |  |
| More than once race | 0 | 1 (0.9%) |  |
| Other/Unknown | 51 (23.8%) | 10 (9.1%) |  |
| **Histologic diagnosis n (%)** |  |  | 0.0005<sup>+</sup> |
| Pilocytic Astrocytoma | 52 (24.2%) | 68 (61.8%) |  |
| Fibrillary Astrocytoma | 0 | 8 (7.3%) |  |
| Pilomyxoid Astrocytoma | 8 (3.7%) | 17 (15.5%) |  |
| Ganglioglioma | 13 (6.1%) | 0 |  |
| Dysembryoplastic neuroepithelial tumor | 7 (3.3%) | 0 |  |
| Diffuse Astrocytoma | 1 (0.5%) | 7 (6.4%) |  |
| Angiocentric Glioma | 1 (0.5%) | 1 (0.9%) |  |
| Other Low-Grade Glioma/Astrocytoma | 132 (61.7%) | 9 (8.2%) |  |
| **BRAF Mutation Status n (%)** |  |  | 0.0005<sup>+</sup> |
| V600E | 50 (23.4%) | 17 (15.2%) |  |
| Fusion | 60 (28.0%) | 60 (53.6%) |  |
| Wildtype | 104 (48.6%) | 35 (31.3%) |  |
| **Tumor Locations n (%)** |  |  | 0.0005<sup>+</sup> |
| Cerebellum/Posterior fossa | 40 (18.7%) | 33 (29.4%) |  |
| Temporal lobe | 43 (20.1%) | 12 (10.7%) |  |
| Frontal Lobe | 22 (10.3%) | 4 (3.6%) |  |
| Suprasellar | 6 (2.8%) | 32 (28.6%) |  |
| Optic Pathway | 8 (3.7%) | 17 (14.9%) |  |
| Brainstem | 7 (3.3%) | 9 (7.9%) |  |
| Thalamus | 15 (7.0%) | 2 (1.8%) |  |
| Ventricles | 14 (6.5%) | 2 (11.4%) |  |
| Others | 59 (27.6%) | 1 (0.9%) |  |

*CBTN: Children Brain Tumor Network. The Kruskal-Wallis rank sum test (\*) was performed for numerical data age to test the statistical significance between age medians. The Fisher's Exact test (+) was performed for categorical data to test the statistical significance differences between CBTN and BCH datasets. A p-value less than 0.05 is statistically significant.*

### TransferX improves deep learning model performance and generalizability

Pipeline with TransferX outperformed the pipeline with classifiers trained by RadImageNet FineTune and training from scratch for BRAF mutational status subtype prediction with composite classification AUC: 0.83 (95% CI 0.71-0.88) and 77% accuracy on internal validation, compared to AUC: 0.74 (95% CI 0.62-0.80) and 73% for training from scratch (Fig. 3B&C) (Table S3). All training approaches, including TransferX, were most accurate at identifying BRAF fusion, followed by wild-type and V600E. However, TransferX was the only approach to maintain AUC >0.80 for all individual subtype classifications (Fig. 3A). With TransferX, the pipeline also exhibits robust performance, with an external AUC of 0.74 (n=28), in the classification of BRAF mutation status, particularly with tumor cases originating from traditionally challenging regions for biopsy such as the optic pathway, thalamus, and brainstem.

**Figure 3.**
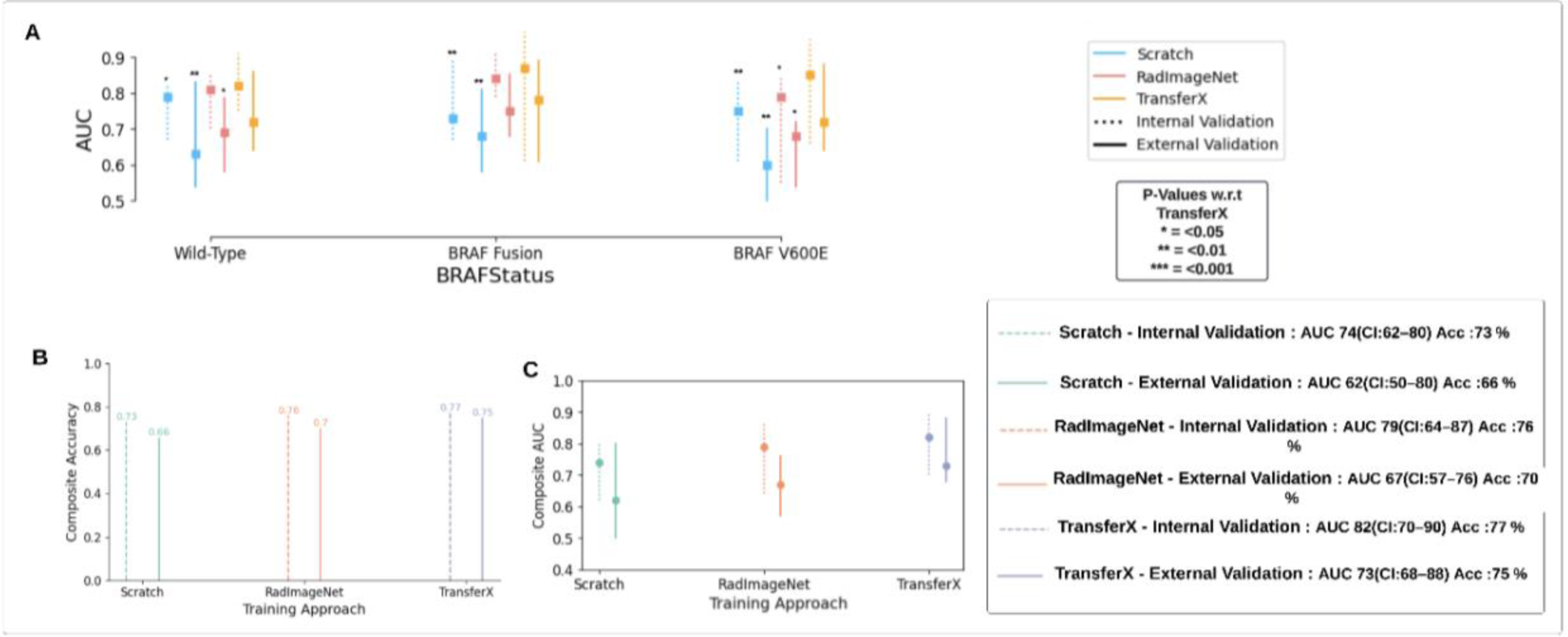
(A) AUC is plotted and compared for the pipeline results with individual subtype classifiers trained using different training approaches (Scratch, RadImageNet FineTune, TransferX) for respective mutation class (BRAF-V600E, BRAF-Fusion, Wild-type). P-values are generated from model comparisons with respect to TransferX. (B) Accuracy and (C) AUC comparison of the pipeline with individual subtype classifiers trained with three different training approaches. The composite Accuracy and AUC for the entire dataset is calculated by the weighted average of the AUCs and Accuracy across the three mutational classes. AUC: area under the curve.

On external validation, there was a mild degradation in performance across all approaches, with TransferX still demonstrating the highest performance with macro-average AUC 0.73 (95% CI: 0.68-0.88) and 75% accuracy (Fig. 3C). TransferX also demonstrated best performance for classification of wildtype vs any BRAF mutational class with AUC 0.82 (95% CI: 0.75-0.91) and 77% accuracy (Table 2 & Fig. 3A). TransferX showed adequate calibration on the external validation set, which was further improved after calibrating the model on the internal validation set (Fig. S7). TransferX also resulted in superior performance compared to other training approaches when subtype classifiers (without consensus logic) were tested on the internal and external validation set for each subtype class (Fig. S6).

**Table 2.** The pipeline’s performance on classification on BRAF status for internal validation set and external validation set.

|  | BRAF Status | AUC (95%CI) | Sensitivity | Specificity | Accuracy | Precision | Recall | F1-Score |
| --- | --- | --- | --- | --- | --- | --- | --- | --- |
| Internal Validation (n=59) | Wild-type | 0.82 (0.75 - 0.91) | 0.73 | 0.80 | 0.77 | 0.76 | 0.77 | 0.77 |
|  | BRAF Fusion | 0.87 (0.61 - 0.97) | 0.87 | 0.70 | 0.81 | 0.81 | 0.80 | 0.80 |
|  | BRAF V600E | 0.85 (0.66 - 0.95) | 0.75 | 0.80 | 0.76 | 0.82 | 0.77 | 0.77 |
|  | Composite | 0.84 (70 - 90) | 0.77 | 0.76 | 0.77 | 0.78 | 0.77 | 0.77 |
| External Validation (n=112) | Wild-type | 0.72 (0.64 - 0.86) | 0.72 | 0.71 | 0.72 | 0.75 | 0.72 | 0.73 |
|  | BRAF Fusion | 0.78 (0.61 - 0.89) | 0.60 | 0.90 | 0.75 | 0.77 | 0.74 | 0.74 |
|  | BRAF V600E | 0.72 (0.64 - 0.88) | 0.78 | 0.60 | 0.75 | 0.82 | 0.74 | 0.77 |
|  | Composite | 0.73 (0.68 - 0.88) | 0.66 | 0.79 | 0.75 | 0.77 | 0.73 | 0.74 |

### TransferX yields more accurate model attention

GradCAMs were generated for the three approaches on all cases (Fig. 4A), and corresponding COMDist scores were calculated. TransferX consistently yielded the best average COMDist scores across all classification tasks, indicating improved model focus on intra- and peritumoral regions (Table 3 & Fig. 4C).

**Figure 4.**
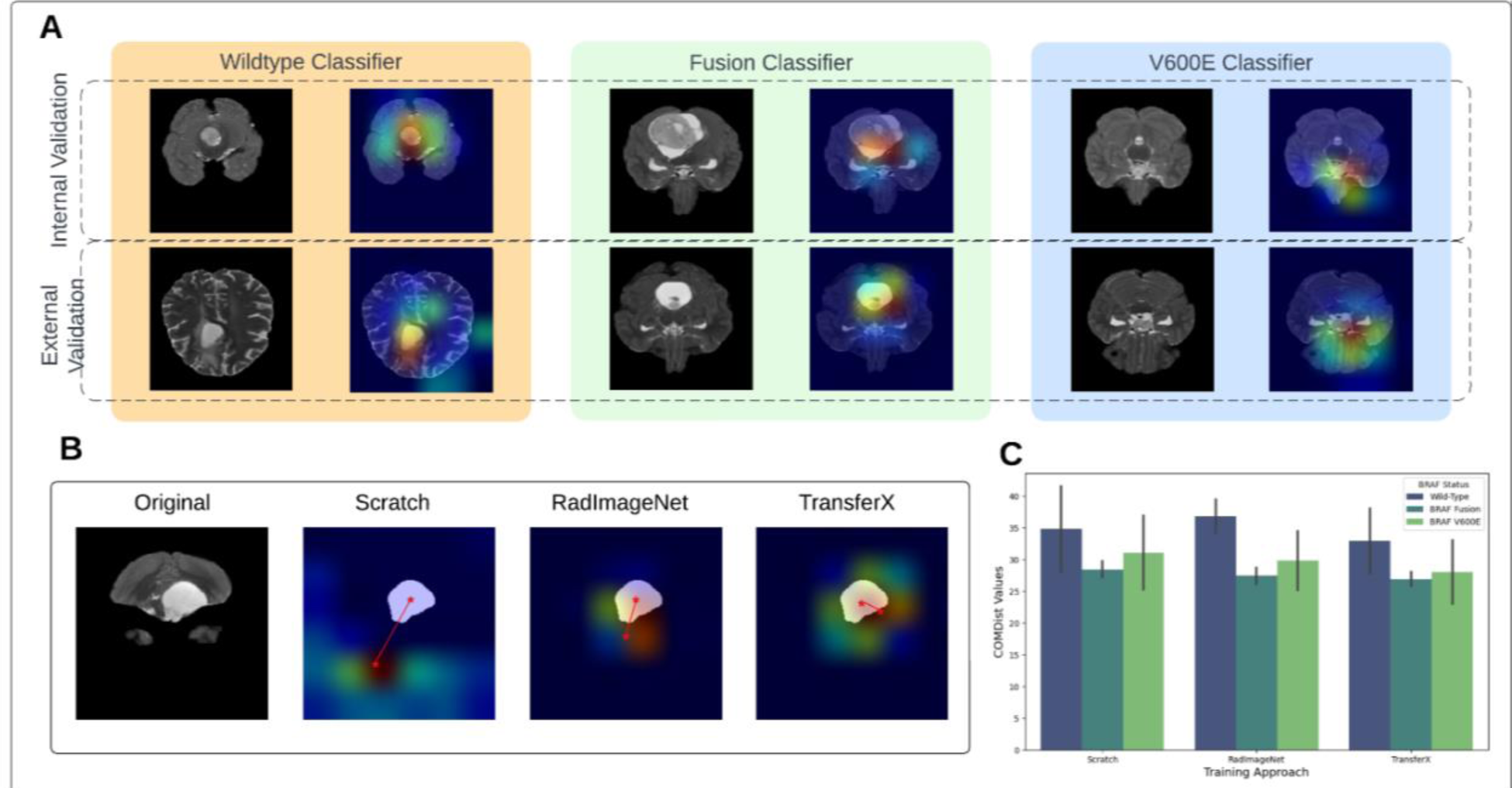
(A) GradCAM image overlay for each mutational class for internal and external validation sets. (B) COMDist representation for three training approaches. (C) COMDist value comparison of the scan-to-prediction pipeline for each molecular subtype class, with corresponding individual subtype classifiers trained with three different training approaches. GradCam: Gradient-weighted Class Activation Maps; COMDist: Center of Mass Distance.

**Figure 5.**
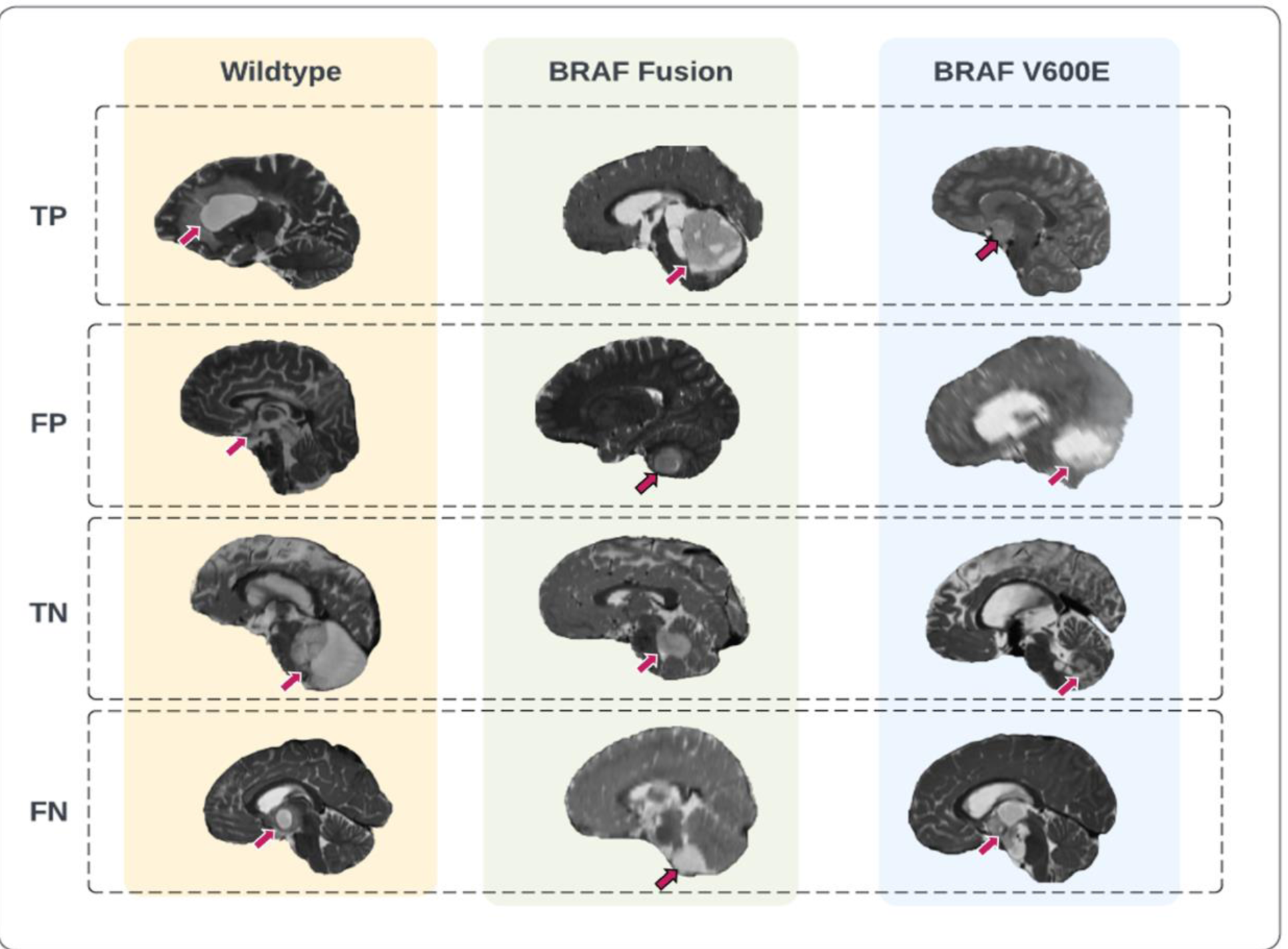
Representative prediction cases of the scan-to-prediction pipeline on the external dataset. The final scan-to-prediction pipeline consists of three subtype classifiers, trained using TransferX, further pooled together in consensus logic by the consensus decision block. Tumor lesions in the T2-weighted images were highlighted with arrows. TP: true positive; FP: false positive; TN: true negative; FN: false negative.

**Table 3.** Median COMDist value (mm) comparison for three training approaches, of each subtype classifier on its corresponding mutation class data.

|  | BRAF Status | TransferX | Scratch | RadImageNet |
| --- | --- | --- | --- | --- |
| Internal Validation (n=59) | Wild-Type | 38.02 | 41.54 (p=0.09) | 39.48 (p=0.46) |
|  | BRAF Fusion | 25.8 | 27.14 (p=0.49) | 26.13 (p=0.86) |
|  | BRAF V600E | 33.02 | 36.86 (p=0.09) | 34.40 (p=0.52) |
| External Validation (n=112) | Wild-Type | 27.8 | 28.11 (p=0.90) | 34.2 (p=0.009) |
|  | BRAF Fusion | 28.0 | 29.7 (p=0.47) | 28.7 (p=0.76) |
|  | BRAF V600E | 23.03 | 25.24 (p=0.40) | 25.21 (p=0.40) |

## DISCUSSION

pLGG can arise in locations that make resection, and even biopsy, morbid and infeasible. In these situations, the ability to noninvasively detect BRAF mutational status via diagnostic imaging would be helpful to determine which patients may benefit from targeted therapies that act on the BRAF pathway and enrollment in clinical trials of novel targeted therapies. In this study, we developed and externally validated a scan-to-prediction algorithm to noninvasively predict BRAF mutational status that could be used in settings where tissue diagnosis is infeasible. The limited quantity of data available for analysis has limited the translational potential of artificial intelligence (AI) in pediatric brain tumor analysis compared to other malignancies. Our study overcomes this obstacle by combining elements of transfer learning and self-supervision to develop a high performing model that maintains good performance on external testing despite heterogeneous tumor and scanner characteristics. Additionally, we introduced COMDist, an interpretable metric to evaluate model attention with anatomic correlation that will help make medical imaging algorithms more trustworthy to clinical users. Our study findings contribute to bridging the gap between AI development and clinical translation in a limited data scenario. To this end, we have published the code and pretrained models to provide usable tools for the scientific community and to encourage clinical testing.

With the emergence of novel BRAF pathway-directed therapies, the segregation of wild-type tumor cases from BRAF subtypes in pLGG has become critical. With an accuracy of ≥77% (Internal) and ≥72% (External) for classifying wild-type tumor cases vs BRAF cases, the pipeline can be used as an assistive tool by clinicians to provide key information in settings where tissue biopsy is infeasible or low-resource settings that preclude genomic analysis. Beyond BRAF classification, the pipeline’s ability to identify BRAF-V600E, specifically, enables it to select patients for specific V600E inhibitors such as of dabrafenib and trametinib which have shown better progression free survival than chemotherapy (29–31). The mild performance degradation observed on external validation may have been driven by notable differences in MRI parameters between across institutions (Fig. S1-2). The model may perform better in scenarios in which MRI parameters are similar to training data. Importantly, the scan-to-prediction pipeline is practical and not reliant on manual segmentation, which is resource-intensive and requires specialized expertise, nor hand-crafted radiomic features, which are notoriously difficult to generalize externally (32–34). Notably, the pipeline also exhibits robust performance in classifying tumors originating from challenging regions for biopsy (optic pathway, thalamus, and brainstem). This may enable more confidence for empiric treatment with targeted therapies if tissue diagnosis is infeasible.

pLGG mutational classification has been previously attempted in a few studies, most with manual segmentation-derived and/or pre-engineered radiomics (35–38), which are known to fail when applied to the external dataset. Radiomic features have been extracted from MRI images and fitted to classifiers models like XGboost and SVM (17,35,36). One study published in preprint, used neural networks to classify BRAF-mutational status in a single institution, though the algorithm required manual segmentation (16). The sensitivity of the dataset size on BRAF mutation classification performance was studied by Wagner et al. in a radiomics based study (39). They showed that Neural networks outperform XGBoost for classification AUC and that the performance was affected by the size of the data used in training. In contrast, our study demonstrates that an end-to-end deep learning pipeline is feasible, even in a low data setting, by using inter-class cross training combined with transfer learning. This idea has been explored more generally by Muhamedrahimov et al. by relaxing the assumption of independence between multiple categories (40). TransferX expands on this work by dropping the assumptions of independence between different categories of a multiclass dataset with stepwise inter-class training as a pretext task to learn robust feature representations. Furthermore, incorporating consensus decision logic to combine multiple binary classifiers also helped mitigate overfitting from the limited dataset.

Interpretability is a well-recognized important factor for deep learning models for clinical translation. A variety of metrics like GradCAM, saliency maps, guided backpropagation have been developed to depict the pixels that are contributing for the maximum activation in the network and hence being more significant for classification (41,42). The GradCAM approach, although adding a degree of qualitative interpretability, has only allowed for case-by-case visualizations for the end-user, which are not very useful when trying to establish trust in a model overall. We expand the utility of GradCAM in this work with COMDist. By incorporating spatial knowledge of the tumor from auto-segmentation, COMDist can quantify, in terms of distance, the model’s attention with respect to the correct, biologically rational region of interest in the image. This provides the clinical user with a metric to gauge whether the model is basing its prediction on intra-tumoral information (as one would expect) or extemporaneous information far from the tumor (indicating an implausible model “shortcut” that should not be trusted). The metric can be reported case-by-case or in aggregate over a dataset to compare attention of different models. We expect this methodology will be valuable for the AI research community as well as clinical end-users evaluating and implementing medical imaging AI applications in clinic.

### Limitations

There are several limitations to this work. Firstly, this work is retrospective in nature and subject to the biases of our patient samples. We attempted to mitigate this effect of bias by using a blinded, external validation set. Thus, we would encourage further independent validation of our results, including prospective testing. Additionally, the pipeline is exclusively based on T2W-MRI scans. While T2W images are the most common and available diagnostic sequence for pLGG, contrast-enhanced T1W, T1W, T2-FLAIR, and diffusion-weighted MRI may contain complementary information that enhances performance. Along with this, the properties of different imaging sequences and their correlation with different molecular subgroups warrants further investigation, which we aim to explore in future work. In this work, we decided to leverage a 2D approach with slice-averaging to minimize overfitting on our limited data set. It is possible that with further data collection a 3D approach may work better, however this would significantly increase the model parameter size and thus make the model even more prone to overfitting.

### Conclusions

In summary, we developed and externally validated an imaging-based scan-to-prediction pipeline to analyze T2W-MRI as input and output BRAF-mutational subtype for pediatric low-grade glioma. We leveraged a novel combination of transfer learning and self-supervision to mitigate overfitting and develop a high-performing and generalizable model. We also proposed a novel evaluation metric, COMDist, that can be used to further assess performance and interpretability of AI imaging models. Our resulting pipeline warrants prospective validation to determine if it could be clinically used in settings where tissue and/or genomic testing is unavailable.

## Author Contributions

Study design: D.T., Z.Y. and B.H.K.; code design, implementation and execution: D.T. and Z.Y.; acquisition, analysis or interpretation of data: D.T., Z.Y., A.Z., and B.H.K.; writing of the manuscript: D.T., Z.Y., B.H.K.; critical revision of the manuscript for important intellectual content: all authors; statistical analysis: Y.Z. and D.T.; study supervision: B.H.K., H.J.W.L.A., T.P., and D.H.K.

## Code availability

The code of the deep learning system, as well as the trained model and statistical analysis are publicly available at the GitHub webpage: https://github.com/DivyanshuTak/BRAF_Classification.

## Data Availability

All data produced in the present study are available upon reasonable request to the authors

https://github.com/DivyanshuTak/BRAF_Classification

