## supplement for "Noninvasive molecular subtyping of pediatric low-grade glioma with self-supervised transfer learning" (from another version of this preprint: version 2, 2023-08-24)

### Supplemental Material

#### Supplementary Methods

#### MR Imaging Acquisition and Parameters

Patients from the Children's Brain Tumor Network (CBTN) cohort underwent brain MR imaging at 1.5T or 3T across various vendors (Siemens; GE Healthcare; Philips Healthcare; Hitachi; Toshiba). Because the CBTN imaging data was collected across several institutions as part of clinical standard of care, images were acquired with non-uniform acquisition protocols. Sequences acquired included 2D axial T2-weighted turbo spin-echo (TR/TE, 3000–7000/90–120 ms; 1- to 5-mm section thickness; 3- to 7.5-mm gap), 2D axial and coronal T2W FLAIR, 3D axial or sagittal pre-contrast, and 3D axial gadolinium-based contrast agent–enhanced T1-weighted turbo or fast-field echo. Patients from the BCH underwent brain MR imaging at 1.5T or 3T from various MRI vendors (Siemens; GE Healthcare). MRIs were performed using the brain tumor protocol of the institution, which included 2D axial T2-weighted fast spin-echo (TR/TE, 7000–10,000/140–170 ms; 4- to 5-mm section thickness; 1- to 1.5-mm gap), 2D axial or sagittal pre-contrast T1-weighted spin- echo, 2D axial T2 FLAIR and 2D axial gadolinium-based contrast agent–enhanced T1-weighted spin-echo sequences. MR acquisition details and parameters for all datasets can be found in Table S1 and Fig. S1-2. All MR imaging data were extracted from the respective PACS and metadata were de-identified for further analyses. Given that many pLGG often do not enhance with intravenous contrast, are hypointense on T1, and are hyperintense on T2-weighted sequence, we chose to develop our algorithm on T2 weighted sequences.

#### MR Image Preprocessing

MRI images were converted from DICOM format to NIFTI format via rasterization packages utilizing dcm2nii package (https://www.nitrc.org/projects/dcm2nii) in Python v3.8. N4 bias filed correction was adopted to correct the low frequency intensity non-uniformity present in MRI images using SimpleITK in Python v3.8. All scans were resampled to 1×1×1 $mm^3$ voxel size using linear interpolation via SimpleITK. After interpolation, the MRI scans were co-registered using a rigid registration step with SimpleITK. Lastly, a brain extraction step was performed for all the scans using HD-BET package (https://github.com/MIC-DKFZ/HD-BET). Imaging acquisition details are found in Supplementary Methods: SM 1.

**Tumor slice selection**

The segmentation mask corresponding to the preprocessed tumor image, outputted by the segmentation model, is employed to extract the top and the bottom slice index (Zmin, Zmax) along the Z axis. The preprocessed image is sliced in axial plane from index ranging from Zmin to Zmax. The remaining slices are discarded as they do not present tumor information. Resulting in the generation of a dataset containing only tumor slices. Both the image and segmentation mask are produced in a dimension of (170, 206, 162), with a voxel spacing of 1mm. The number of 2D tumor slices generated from a single 3D scan is dependent on the tumor length along the Z-axis. The resulting 2D images are further normalized and resized to a shape of (192, 192).

Prior to slice extraction, segmentation model output is post-processed to remove small islands below a volume of 500 $mm^3$, which may represent artifact or small satellite lesions. For the slicing and extraction of 2D tumor images, the axial plane is selected, given that this is how MRI brain data is most commonly acquired, including our publicly available MRI imaging datasets including the RadImageNet dataset. This allows us to use the publicly available pretrained model weights for training of the individual subtype classifiers.

**Deep learning training details**

Each subtype classifier has a ResNet50 encoder backbone, which is extracted before the average pool layer. The backbone encoder network was further appended by two fully connected layers with 1024 neurons in the penultimate layer and a single neuron in the final layer to perform binary classification. The entire pipeline was implemented in Tensorflow V2.0, with additional Python libraries including nibabel and pillow were used to facilitate pre and post processing on the dataset. All the training were done on an Nvidia A6000 GPU.

The developmental dataset was split into a train and validation set for the experiments, with a split ratio of 75:25 (Fig. S4). Data augmentations like gaussian blur, random rotation (probability = 0.5, maximum angle = 10), horizontal flip, and resampling with nearest neighbor interpolation were used to enable robust model training and prevent overfitting. A batch size of 32 was used for all the training and fine tuning with a constant learning rate of 1e-4. Each training and fine-tuning step was performed for 50 epochs. A binary cross entropy loss function coupled with sigmoid activation for the last layer was used for loss calculation during training. Each subtype classifier consists of a ResNet50 backbone and is trained with a constant input size of (192,192,3).

During the sequential training, for TransferX, early convolutional layers were not frozen and the whole network was fine-tuned end-to-end. For the RadImageNet FineTune method, the pretrained weights were loaded for the convolutional layers, before the average pooling layer, and the final classification layers were initialized with random weights. Training from scratch involved randomly initializing the weights of the entire network prior to training.

**Subtype Classifier**

Three individual binary subtype classifiers were trained, wild-type classifier, BRAF Fusion classifier, and BRAF V600E classifier. For the training, the multi-class development dataset, with instances of wild-type, BRAF fusion and BRAF V600E, was divided into three binary datasets in a “one Vs rest” format. Each subtype classifier was trained and inferred on the corresponding One Vs rest dataset. For example, wildtype classifier was trained and inferred on the wild-type Vs rest binary dataset, similarly for the other subtype classifiers. The external validation dataset was split into three one Vs rest dataset for external validation of the individual subtype classifiers and the entire pipeline.

**Consensus decision logic**

We hypothesized that morphologic differences (and signal-to-noise ratio) between wild-type and any mutations are greater than between BRAF mutation subtypes, thus wild-type mutation check is performed first. In instances where the patient exhibits a wild-type mutation, signifying the absence of a BRAF mutation, the diagnostic process culminates. Conversely, if the patient possesses any BRAF mutation, further classification between BRAF-Fusion and BRAF-V600E mutations is performed. In this way, the use of sequential logic and binary classification form a rationale path to overall mutational status prediction and avoids the need for multi-class algorithms that would increase the risk of overfitting on a limited dataset.

The principles of consensus logic, as detailed by the following mathematical formulations, are based upon classifications and its corresponding score from each individual subtype classifiers. We designate 'A' as the wild-type classifier, 'B' as the fusion classifier, and 'C' as the v600e classifier. Additionally, we represent the event where the wild-type classifier (A) predicts an instance as belonging to the wild-type class (A=1) by 'α'. Similarly, 'β' and 'γ' respectively denote the events where the fusion classifier (B) identifies an instance as a fusion class and the v600e classifier (C) classifies an output as belonging to the v600e class.

In the same tangent, we introduce 'ε', 'φ', and 'ꞷ' to represent decisions outputted by the consensus logic. Specifically, 'ε' represents the event in which consensus logic predicts the output as belonging to the wild-type class, 'φ' signifies the event of a output is predicted as a fusion class, and 'ꞷ' denotes the event when the output is classified as v600e class. $\tilde{\beta}$ represents the scenario where B = 0, and $\tilde{\gamma}$ represents the case when C = 0. P( ) denotes the associated probability score of the encompassed event.

$$\varepsilon = \alpha \cap (\tilde{\beta} \cup \tilde{\gamma}) \quad (1)$$

$$P(\varepsilon) = P(\alpha) \cap (P(\tilde{\beta}) \cup P(\tilde{\gamma}))$$

$$P(\varepsilon) = P(\alpha) * (P(\tilde{\beta}) + P(\tilde{\gamma}) - P(\tilde{\beta}) * P(\tilde{\gamma})) \quad (2)$$

$$\tilde{\varepsilon} = \sim(\alpha \cap (\tilde{\beta} \cup \tilde{\gamma}))$$

$$P(\tilde{\varepsilon}) = 1 - P(\varepsilon)$$

$$\varphi = \beta \cap \tilde{\varepsilon}$$

$$\varphi = \beta \cap (\sim(\alpha \cap (\tilde{\beta} \cup \tilde{\gamma}))) \quad (3)$$

$$P(\varphi) = P(\beta) * P(\tilde{\varepsilon})$$

$$P(\varphi) = P(\beta) * (1 - (P(\alpha) * (P(\tilde{\beta}) + P(\tilde{\gamma}) - P(\tilde{\beta}) * P(\tilde{\gamma})))) \quad (4)$$

$$\tilde{\varphi} = \sim(\beta \cap \tilde{\varepsilon})$$

$$P(\tilde{\varphi}) = 1 - P(\varphi)$$

$$\omega = \gamma \cap \tilde{\varphi}$$

$$\omega = \gamma \cap (\sim(\beta \cap (\sim(\alpha \cap (\tilde{\beta} \cup \tilde{\gamma}))))) \quad (5)$$

$$P(\omega) = P(\gamma) * (1 - P(\beta) * (1 - (P(\alpha) * (P(\tilde{\beta}) + P(\tilde{\gamma}) - P(\tilde{\beta}) * P(\tilde{\gamma}))))) \quad (6)$$

Each equation elucidates the way outputs from individual subtype classifiers coalesce within the consensus logic to predict the final pipeline outcomes. Specifically, Equation (1) sets forth the consensus logic's classification output for the wild-type class, with its corresponding probability score detailed in Equation (2). This forms the first check in the sequence of decision-making.

If the input is not classified as Wild-type, the consensus logic progresses to consider the possibility of a fusion classification. This event is characterized in Equation (3), and its corresponding probability score is illustrated in Equation (4).

If the input does not match either the Wild-type or Fusion classifications, the consensus logic then explores the possibility of a v600e classification. Equation (5) thus interprets this event of consensus logic predicting an output as v600e, while Equation (6) calculates its corresponding probability score. This forms the final step in the classification process, ensuring the examination of all potential classification outcomes.

**Model calibration**

The subtype classifiers (Wild-type, Fusion, and V600E) were calibrated using the Scikit-Learn implementation of CalibratedClassifierCV. This method calibrates the classifier with probability calibration via isotonic or sigmoid regression. Importantly, the calibration process was conducted using an internal dataset, which was partitioned into a 70:30 split. Here, 70% of the data was used for the calibration training set, and the remaining 30% was used as calibration validation set for the calibration process. The trained calibration was then applied on the external set (Fig. S7).

**Supplementary Tables**

**Table S1.** MRI Machine distribution for developmental dataset.

| Manufacturer | Model | Patients | Percentage (%) |
|---|---|---|---|
| GE Medical Systems | GENESIS_SIGNA | 50 | 21.18 |
| GE Medical Systems | Optima MR450w | 1 | 0.42 |
| GE Medical Systems | SIGNA EXCITE | 47 | 19.91 |

| | | | |
|---|---|---|---|
| GE Medical Systems | SIGNA HDx | 5 | 2.11 |
| GE Medical Systems | Signa HDxt | 22 | 9.32 |
| HITACHI Medical Corporation | Altaire | 1 | 0.42 |
| Philips Healthcare | Ingenia | 1 | 0.42 |
| Philips Medical Systems | Achieva | 1 | 0.42 |
| Philips Medical Systems | Intera | 3 | 1.27 |
| Philips Medical Systems | Panorama HFO | 1 | 0.42 |
| SIEMENS | Avanto | 5 | 2.11 |
| SIEMENS | Espree | 8 | 3.38 |
| SIEMENS | Skyra | 25 | 10.59 |
| SIEMENS | Sonata | 1 | 0.42 |
| SIEMENS | Symphony | 19 | 8.05 |
| SIEMENS | TrioTim | 41 | 17.37 |
| SIEMENS | Verio | 4 | 1.69 |
| TOSHIBA | Titan | 1 | 0.42 |

**Table S2.** P-value comparison of the pipeline, with individual subtype classifiers trained with 3 different approaches followed by the consensus logic. The comparison is performed with respect to TransferX.

| | BRAF Status | Scratch | RadImageNet |
|---|---|---|---|
| Internal Validation (n=59) | Wild-Type | 0.04 | 0.87 |
| | BRAF Fusion | 0.002 | 0.35 |
| | BRAF V600E | 0.003 | 0.04 |
| External Validation (n=112) | Wild-Type | 0.005 | 0.02 |
| | BRAF Fusion | 0.005 | 0.66 |
| | BRAF V600E | 0.007 | 0.05 |

**Table S3.** P-value comparison of individual subtype classifiers for corresponding subtype class. TransferX with training from scratch and RadImageNet finetune for each subtype classifier.

| | BRAF Status | Scratch | RadImageNet |
|---|---|---|---|
| Internal Validation (n=59) | Wild-Type | 0.03 | 0.04 |
| | BRAF Fusion | 0.004 | 0.29 |
| | BRAF V600E | 0.02 | 0.33 |
| External Validation (n=112) | Wild-Type | 0.009 | 0.02 |
| | BRAF Fusion | 0.007 | 0.72 |
| | BRAF V600E | 0.0003 | 0.01 |

**Table S4.** Instance count of variables age, sex, tumor location for each subtype class. For categorical variables of sex and tumor location a Chi-Squrared test was performed to test the statistically significant differences among molecular subtypes.

| | Variable | BRAF V600E | BRAF Fusion | Wild-type | P-value |
|---|---|---|---|---|---|
| **Sex** | Male | 27 | 28 | 40 | 0.71 |
| | Female | 26 | 32 | 55 | |
| | Other | 3 | 1 | 2 | |
| **Tumor Location** | Brainstem | 2 | 3 | 2 | 0.0002 |
| | Frontal lobe | 2 | 5 | 15 | |
| | Optic pathway | 3 | 3 | 0 | |
| | Cerebellum | 5 | 22 | 13 | |
| | Suprasellar | 2 | 1 | 3 | |
| | Temporal lobe | 20 | 5 | 18 | |
| | Thalamus | 3 | 4 | 6 | |
| | Ventricle | 2 | 5 | 4 | |
| | Other | 16 | 12 | 38 | |

### Supplementary Figures

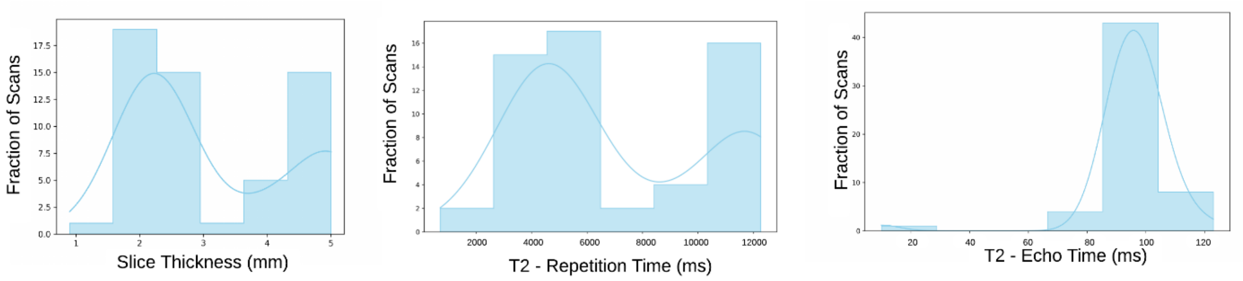


**Figure S1.** Slice thickness, Repetition time and Echo time for T2 Weighted MRI Images for Developmental Dataset (BCH).

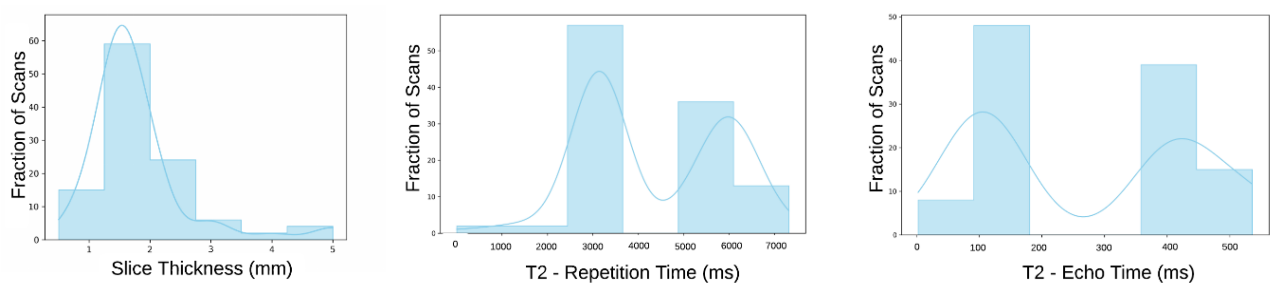


**Figure S2.** Slice thickness, repetition time and echo time for T2 Weighted MRI Images for the external validation dataset (CBTN).

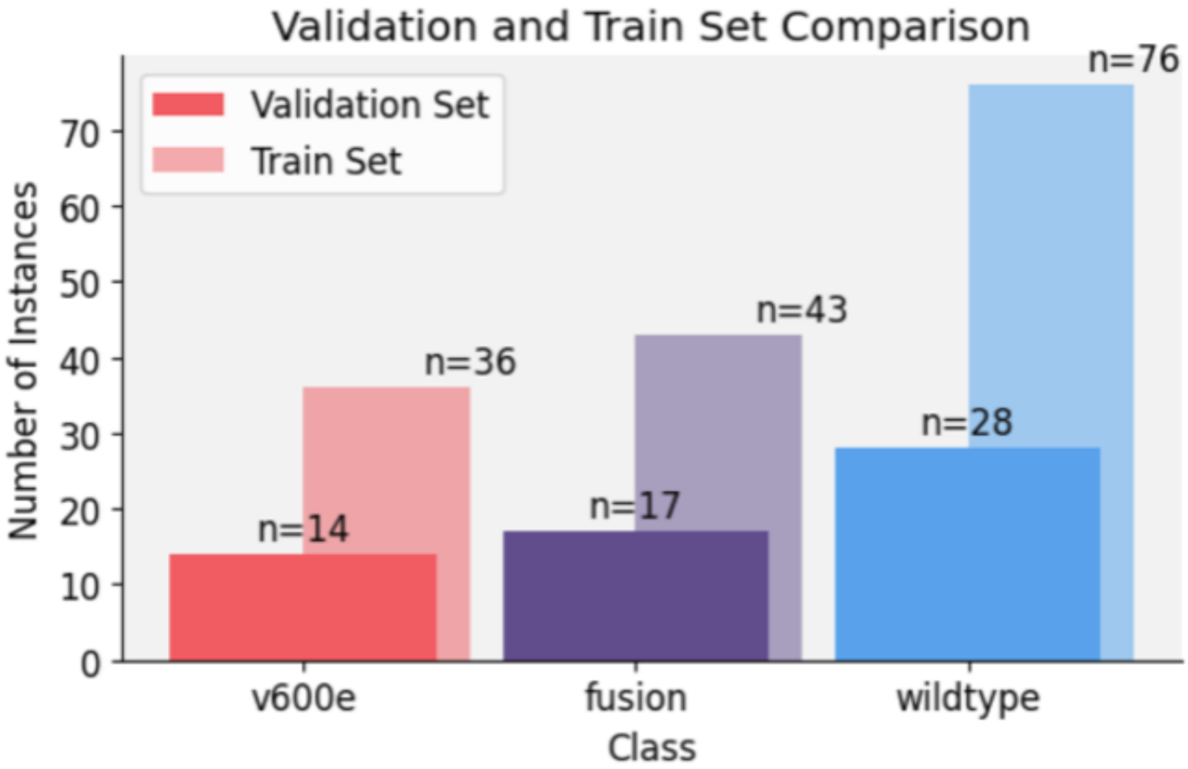


**Figure S3.** Distribution details for training and validation split for developmental dataset.

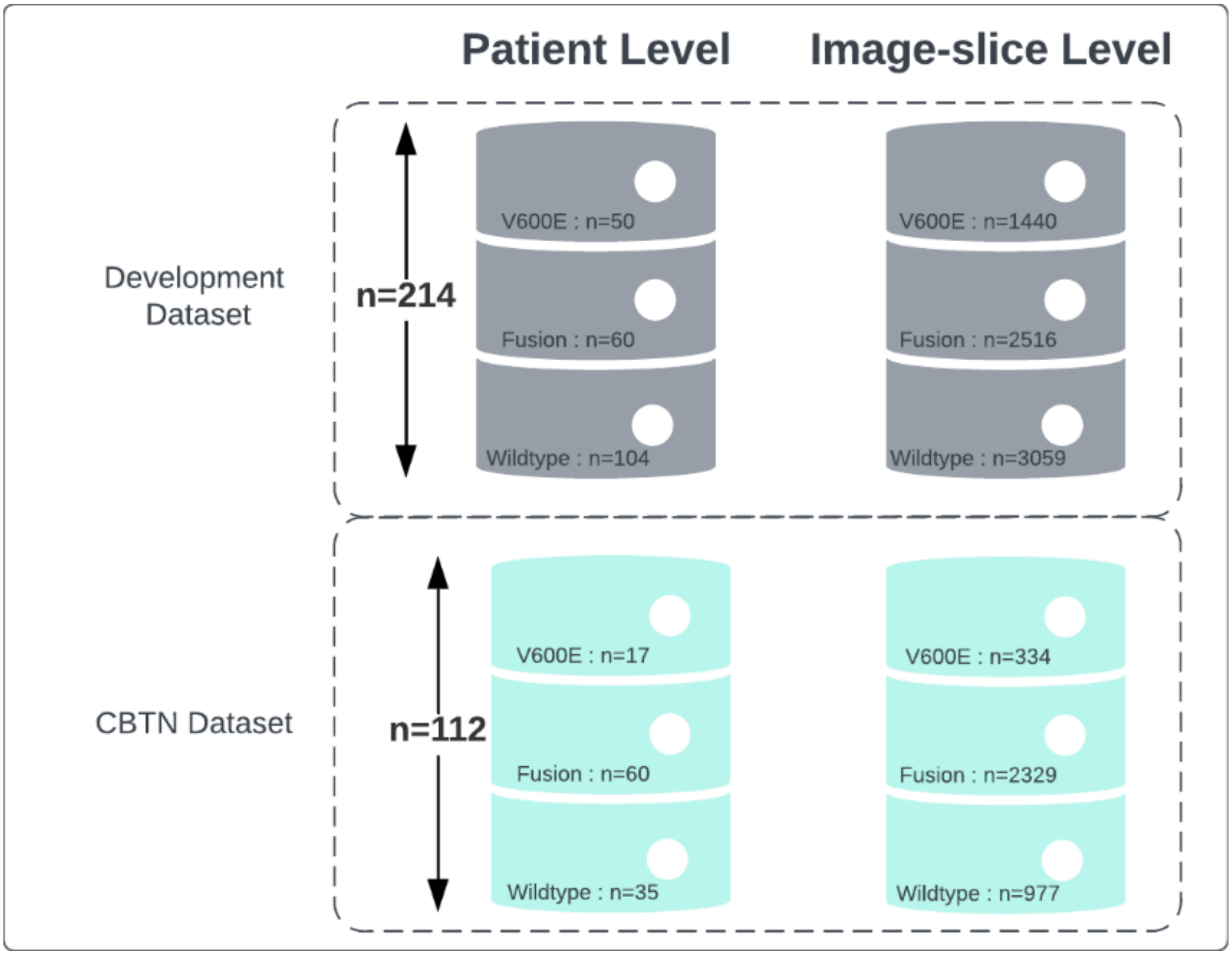


**Figure S4.** Patient-level and Image-slice level instance distribution of the developmental dataset and CBTN dataset used for the study.

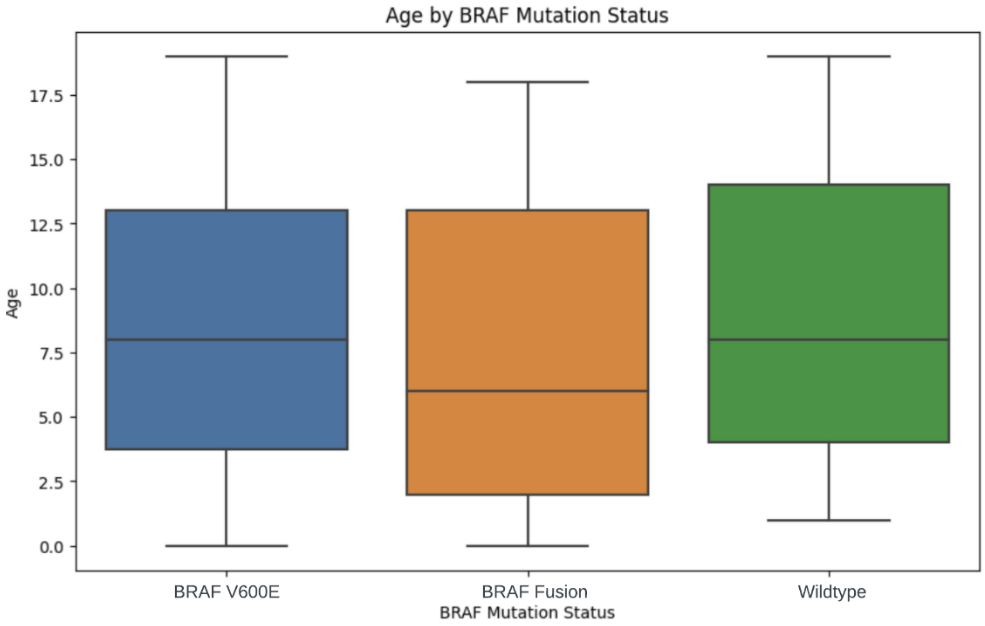


**Figure S5.** Distribution of Age with respect to BRAF Mutation status for BCH dataset. One-way ANOVA test was performed to check the correlation with BRAF Mutational status (P=0.14).

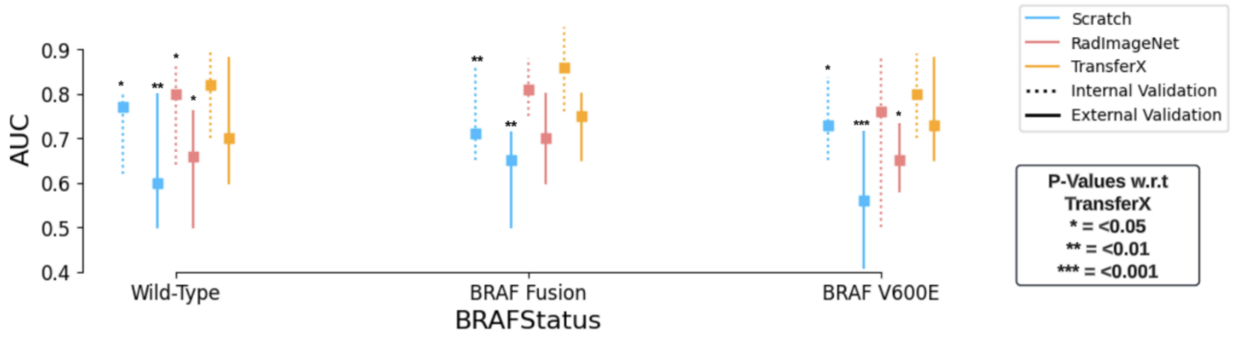


**Figure S6.** AUC is plotted and compared for each individual molecular subtype classifier for different training approaches (Scratch, RadImageNet FineTune, TransferX) for respective mutation class (wild-type, BRAF fusion, BRAF V600E). P-values are generated from model comparisons with respect to TransferX.

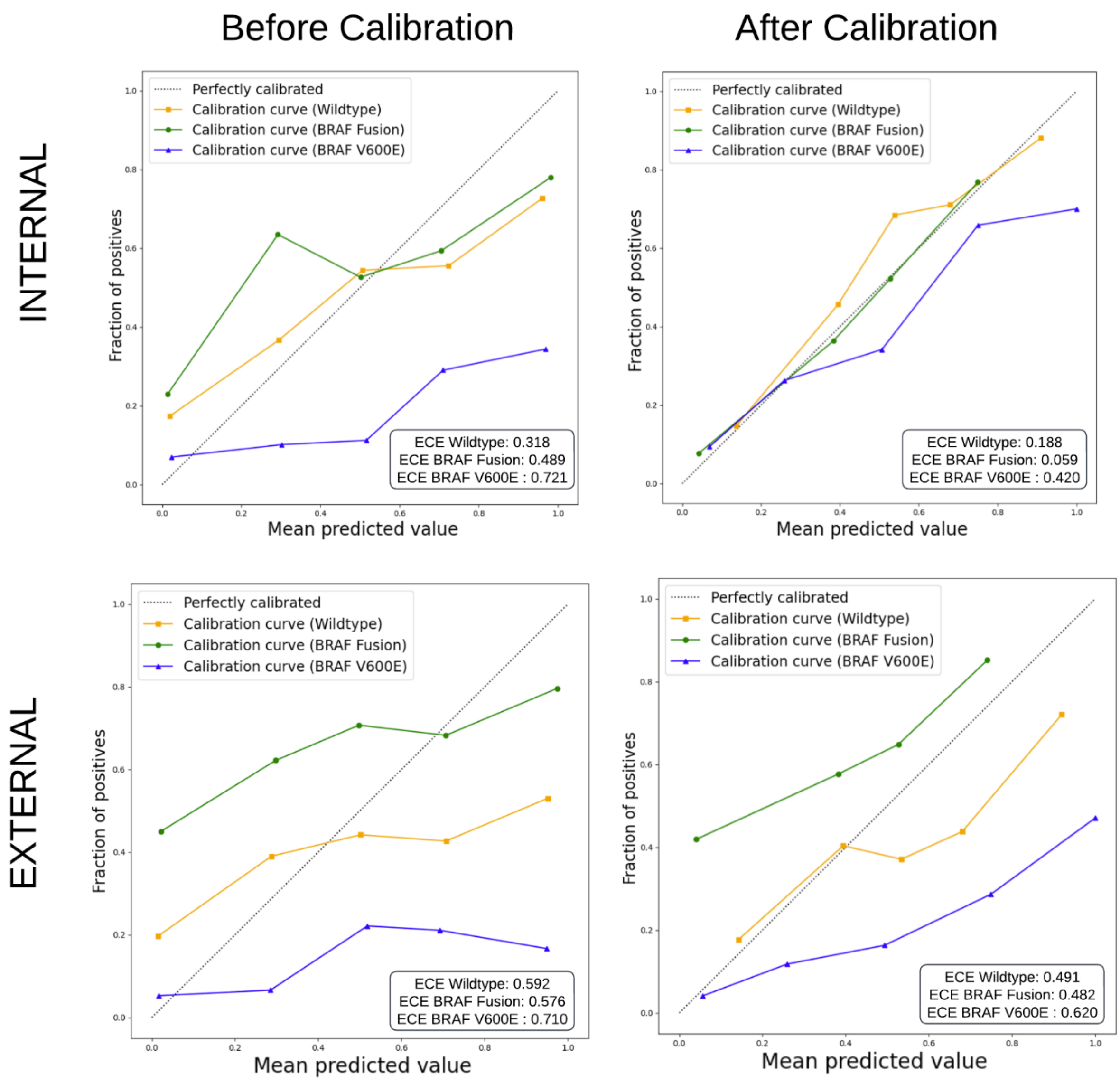


**Figure S7.** Calibration Curves for individual subtype classifiers for developmental and CBTN dataset.
